# Multi-ancestry GWAS analysis identifies two novel loci associated with diabetic eye disease and highlights *APOL1* as a high risk locus in patients with diabetic macular edema

**DOI:** 10.1101/2023.01.23.23284446

**Authors:** Amy D. Stockwell, Michael C. Chang, Anubha Mahajan, William Forrest, Neha Anegondi, RK Pendergrass, Suresh Selvaraj, Jens Reeder, Eric Wei, VA Iglesias, Natalie M. Creps, Laura Macri, Andrea N. Neeranjan, Marcel P. van der Brug, Suzie J. Scales, Mark I. McCarthy, Brian L. Yaspan

## Abstract

Diabetic retinopathy (DR) is a common complication of diabetes. Approximately 20% of DR patients have diabetic macular edema (DME) characterized by fluid leakage into the retina. There is a genetic component to DR and DME risk, but few replicable loci. Because not all DR cases have DME, we focused on DME to increase power, and conducted a multi-ancestry GWAS to assess DME risk in a total of 1,502 DME patients and 5,603 non-DME controls in discovery and replication datasets. Two loci reached GWAS significance (p<5×10^−8^). The strongest association was rs2239785, (K150E) in *APOL1*. The second finding was rs10402468, which co-localized to *PLVAP* and *ANKLE1* in vascular / endothelium tissues. We conducted multiple sensitivity analyses to establish that the associations were specific to DME status and did not reflect diabetes status or other diabetic complications. Here we report two novel loci for risk of DME which replicated in multiple clinical trial and biobank derived datasets. One of these loci, containing the gene *APOL1*, is a risk factor in African American DME and DKD patients, indicating that this locus plays a broader role in diabetic complications for multiple ancestries.

## Introduction

Diabetic retinopathy (DR) is a common complication of diabetes and one of the leading causes of blindness worldwide(1). Approximately 20% of DR patients have diabetic macular edema (DME) characterized by fluid leakage into the retina(2). DME can occur at any point in the timeline of DR, but is more likely to occur later in the disease. Diabetic patients with African American ancestry have the greatest risk of developing DME of all populations in the United States, estimated at three times greater than those of European-descent(3). Risk factors include improper glycemic control and duration of diabetes, but these do not explain all of the heterogeneity in DME risk (4-6). While it is accepted there is a genetic component to DR and DME risk, to date, large-scale genome-wide association studies (GWAS) have not yielded robust insights(7). Most well powered studies have been in DR, and have been well summarized(8). DR is an inherently heterogeneous disease marker, and one suggestion cited for identification of replicable loci identified via GWAS was harmonized patient phenotyping (9). As an example, one recent study containing DR, DME and proliferative diabetic retinopathy cases in a multi-ethnic study population of 43,565 was unable to find loci at genome-wide significance (10). Because not all DR cases have DME, focusing on that subtype alone could increase power to find novel loci associated with the risk of disease. Furthermore, the increased prevalence of DR and DME in African American individuals indicates there may be genetic risk factors specific to that population. To that end, we performed the largest genetic study of DME to date (a total of 1,502 cases and 5,603 controls) identifying two loci reaching genome-wide significance for risk of DME, one of which contains *APOL1* and is estimated to account for approximately 5% of the risk of developing DME in African American patients with diabetes.

## Methods

### Study Design and Populations

We performed a genome wide association study study using DNA derived from blood samples obtained from DME patients from clinical trials for ranibizumab (NCT00473330 [RISE] and NCT00473382 [RIDE]), faricimab (NCT03622580 [YOSEMITE] and NCT03622593 [RHINE]), and the port delivery system with ranibizumab (NCT04108156 [PAGODA]). These study populations were selected for inclusion on the basis of available phenotypic information and DNA availability for whole-genome sequencing (all but PAGODA) and array genotyping (PAGODA). Patients were sequenced and genotyped between 2017-2021.

Samples and data for controls in the discovery study population (cases from the ranibizumab (RISE and RIDE) and faricimab (YOSEMITE and RHINE) studies) were obtained from clinical trial studies of asthma, autoimmune disease, Crohn’s disease, chronic obstructive pulmonary disease, inflammatory bowel disease, interstitial lung disease, idiopathic pulmonary fibrosis, rheumatic arthritis, systemic lupus erythematosus and ulcerative colitis. An independent set of controls for the replication study population (cases from the port delivery study with ranibizumab – PAGODA) were selected from clinical trial studies of cancer and asthma, in addition to healthy controls from a cohort study examining Age-Related Macular Degeneration in subjects with African ancestry (DIVERSITY).

All patients included in this study provided written informed consent for whole-genome sequencing or array genotyping of their DNA. Ethical approval was provided as per the original clinical trials.

### DNA analysis

The WGS data for the discovery study population was generated to a read depth of 30X using the HiSeq platform (Illumina X10, San Diego, CA, USA) processed using the Burrows-Wheeler Aligner (BWA) / Genome Analysis Toolkit (GATK) best practices pipeline. WGS short reads were mapped to hg38 / GRCh38 (GCA_000001405.15), including alternate assemblies, using BWA version 0.7.9a-r786 to generate BAM files. All WGS data was subject to quality control and checked for concordance with SNP fingerprint data collected before sequencing. After filtering for genotypes with a GATK genotype quality greater than 90, samples with heterozygote concordance with SNP chip data of less than 75% were removed. Sample contamination was determined with VerifyBamID software, and samples with a freemix parameter of more than 0.03 were excluded. Joint variant calling was done using the GATK best practices joint genotyping pipeline to generate a single variant call format (VCF) file. The called variants were then processed using ASDPEx to filter out spurious variant calls in the alternate regions.

Data in the replication study population was generated using the Infinium Global Screening Array-24 Kit v2 and v3 from Illumina. Genotypes with GenCall (GC) score less than 0.15 were set to missing.

### Quality Control – Patient DNA Samples

Ancestry was assigned using an ancestry threshold of 0.7 obtained from supervised analysis with ADMIXUTRE 1.3 using samples from phase 3 of the 1000 Genomes Project as reference samples labeled by their superpopulation (**Supplemental Figure 1**) (AFR = African [from the African continent and African ancestry from the US and Caribbean]; AMR = AdMixed American [from North and South America excluding US of African Ancestry and Caribbean of African ancestry]; EAS = East Asian [from China, Japan and Vietnam]; EUR = European [from the United States with Northern and Western European ancestry, Italy, Finland, England and Scotland]).

For the discovery study population, MatchIt(11) was used to do a 4:1 (control:case) ratio using the nearest neighbor matching on the propensity score to select controls based on age, sex and global ancestry.

For both the discovery and replication study populations, samples were excluded if the call rate was less than 90%. Identity by descent analysis was used to detect and filter out relatedness in the dataset; samples were excluded if PI_HAT was 0.4 or higher. Samples were removed if they showed excess heterozygosity with more than three SDs of the mean. This resulted in 448 in the AFR study population (99 cases and 349 controls), 343 in the AMR study population (87 cases and 347 controls), 439 in the EAS study population (88 cases and 351 controls) and 4,577 in the EUR study population (921 cases and 3,656 controls) for the discovery dataset with a combined total of 1,195 DME cases and 4,703 non-DME controls. Similarly, after quality control there are 976 patient samples in the EUR replication study population (262 cases and 714 controls) and 231 subjects in the AFR replication study population (45 cases and 186 controls) for a combined total of 307 DME cases and 900 non-DME controls.

### Quality Control - Genotype

For the replication study population, multi-allelic sites were split into biallelic records, variants were left-aligned and normalized, and duplicate variants based on chromosome, position, ref allele, and alt allele were removed prioritizing by missingness rate. Additional quality control was performed by removing variants that had a target majority ancestry alternate allele frequency (AF) difference exceeding 0.2 compared to the reference, as well as removing ambiguous (A/T, G/C) variants where target majority ancestry minor allele frequency (MAF) exceeded 0.4 in either the target or reference. Phasing and imputation were carried out using Beagle 5.1 with hg38 HapMap genetic maps and a reference panel consisting of merged Genentech internal whole-genome sequencing and Haplotype Reference Consortium (HRC) genotypes. This merged panel consists of 134,246,541 bi-allelic sites in 51,455 (chr1), 55,929 (chr2-22), and 54,963 (chrX) samples. The default sliding window length of 40 cM with 4 cM overlap between adjacent sliding windows was used. Imputed VCFs were annotated with rsIDs from dbSNP154. The estimated squared correlation between the estimated allele dose and the true allele dose (DR2) was used to filter out poorly imputed variants by setting a DR2 threshold of 0.3.

Sample genotypes (discovery WGS only) were set to missing if the Genotype Quality score was less than 20 and SNPs were removed if the missingness was higher than 5%. SNPs were filtered if the significance level for the Hardy-Weinberg equilibrium test was less than 5×10^−8^. The allele depth balance test was performed to test for equal allele depth at heterozygote carriers using a binomial test (discovery WGS only); SNPs were excluded if the p-value was less than 1×10^−5^.

### Statistical Analysis

A common variant (MAF>=1%) genome-wide association study (GWAS) was conducted to assess DME risk in the AFR, AMR, EAS and EUR study populations separately. PLINK(12) was used to perform logistic regression two-sided test using an additive model, adjusted for age, sex and genetic ancestry. The first three PCAs were used in as covariates for genetic ancestry and their cumulative proportion of variance was >90%. METAL(13) was subsequently used to do a meta-analysis, under a mixed effects model. There were 7,601,242 variants that passed QC in one or more of the cohorts. The ancestry-specific cohorts were QC’d and analyzed separately, rather than filtering on MAF in the combined cohort. For the meta-analysis, variants were included if they were present in two or more of the ancestry groups.

LDSC software was used separately on EUR and AFR GWAS summary stats to calculate heritability(14). The parp function in the CRAN R library twoxtwo was used to calculate the population attributable risk proportion in EUR and AFR study populations combined and separately(15).

Samples from other disease areas were used as controls. We used a genotype-on-phenotype reverse regression to test for association with the non-DME controls. We calculated the posterior probability (PP) that a SNP is also associated with each individual disease area included in the control cohort; SNPs passing the 0.6 PP threshold in any of the control cohort disease areas were flagged and excluded from the GWAS results.

### FinnGen ethics statement and methods

A second replication population consisted of patients enrolled in the FinnGen Biobank cohort. Patients and control subjects in FinnGen provided informed consent for biobank research, based on the Finnish Biobank Act. Alternatively, separate research cohorts, collected prior the Finnish Biobank Act came into effect (in September 2013) and start of FinnGen (August 2017), were collected based on study-specific consents and later transferred to the Finnish biobanks after approval by Fimea (Finnish Medicines Agency), the National Supervisory Authority for Welfare and Health. Recruitment protocols followed the biobank protocols approved by Fimea. The Coordinating Ethics Committee of the Hospital District of Helsinki and Uusimaa (HUS) statement number for the FinnGen study is Nr HUS/990/2017.

The FinnGen study is approved by Finnish Institute for Health and Welfare (permit numbers: THL/2031/6.02.00/2017, THL/1101/5.05.00/2017, THL/341/6.02.00/2018, THL/2222/6.02.00/2018, THL/283/6.02.00/2019, THL/1721/5.05.00/2019, THL/1524/5.05.00/2020, and THL/2364/14.02/2020), Digital and population data service agency (permit numbers: VRK43431/2017-3, VRK/6909/2018-3, VRK/4415/2019-3), the Social Insurance Institution (permit numbers: KELA 58/522/2017, KELA 131/522/2018, KELA 70/522/2019, KELA 98/522/2019, KELA 138/522/2019, KELA 2/522/2020, KELA 16/522/2020, Findata THL/2364/14.02/2020 and Statistics Finland (permit numbers: TK-53-1041-17 and TK/143/07.03.00/2020 (earlier TK-53-90-20).

The Biobank Access Decisions for FinnGen samples and data utilized in FinnGen Data Freeze 7 include: THL Biobank BB2017_55, BB2017_111, BB2018_19, BB_2018_34, BB_2018_67, BB2018_71, BB2019_7, BB2019_8, BB2019_26, BB2020_1, Finnish Red Cross Blood Service Biobank 7.12.2017, Helsinki Biobank HUS/359/2017, Auria Biobank AB17-5154 and amendment #1 (August 17 2020), Biobank Borealis of Northern Finland_2017_1013, Biobank of Eastern Finland 1186/2018 and amendment 22 § /2020, Finnish Clinical Biobank Tampere MH0004 and amendments (21.02.2020 & 06.10.2020), Central Finland Biobank 1-2017, and Terveystalo Biobank STB 2018001.

### Colocalization

We utilized colocalization analysis between eQTL data from GTEx, eyeQTL, and Sun et al pqtls to identify putative causal genes in the region(16-18). The coloc library (CRAN R) uses approximate Bayes factors to estimate posterior probabilities (PP) for common variants in the GWAS study as well as the QTL study. A PP4 estimates the probability that a variant is associated with both traits and a PP4 threshold of 0.8 was used as evidence for colocalization. Variants within +/- 250 bp of the most significant SNP in the locus were included. GTEx tissues relevant to complications of diabetes were included: kidney and vascular / endothelium (artery aorta, artery coronary and artery tibial). EyeQTL included the following tissues: RPE, retina, RPE macula, RPE non-macula, retina macula, and retina non-macula.

## Results

We conducted a multi-ancestry GWAS to identify risk alleles associated with DME. Discovery population cases (N=1,195) comprised patients from clinical trials for DME involving ranibizumab (NCT00473330 [RISE N=126], NCT00473382 [RIDE N=143]) and faricimab (NCT03622580 [YOSEMITE N=511] and NCT03622593 [RHINE N=415]) who consented for genetic analysis. Inclusion criteria for all studies allowed for both T1D and T2D patients. Based on self-reported data, from YOSEMITE and RHINE, we estimate 6% of participants have T1D and 94% have T2D. We obtained controls (N=4,703) from participants in non-ophthalmic clinical trials that were sequenced and processed with the same informatics pipeline; this ensures we minimize batch effects that may be introduced via differing sequencing technologies and bioinformatics processing pipelines (see **methods** for details). We grouped DME cases and non-DME controls by genetic ancestry using defined populations in the 1000 Genomes Project. Therefore, population subsets in this study will be referred to by the 1000 Genomes super-population codes (see **methods** for details). We performed case and control analyses for each of the four super-populations (AFR - African, EUR - Euroepan, EAS – East Asian and AMR – AdMixed American), controlling for age, sex and genetically defined ancestry, combining results via meta-analysis. Due to the use of individuals with other, non-ophthalmic diseases as controls, we used a genotype-on-phenotype reverse regression to remove non-DME specific findings, which have been removed from the presented analysis(19).We sought to replicate any genome-wide significant signals in a sample of 307 multi-ancestry DME cases from a clinical trial evaluating ranibizumab in the Port Delivery System (NCT04108156 [PAGODA]) and 900 non-DME controls obtained in a similar fashion to the discovery dataset. Case and control demographics for the discovery and replication populations are available in **Table 1**.

**Table 1.** Cohort Demographics.

|  | Discovery<br>Cases | Replication<br>Controls | P-value | Cases | Controls | P-value |
| --- | --- | --- | --- | --- | --- | --- |
| **AFR** |  |  |  |  |  |  |
| N | 99 | 349 |  | 45 | 186 |  |
| Age, mean (sd) | 61.2 (9.6) | 56.8 (8.6) | 4.10E-05 | 61.1 (8.6) | 71.7 (8.1) | 5.50E-07 |
| Female, N (%) | 62 (62.6%) | 247 (70.7%) | 0.15 | 22 (48.8%) | 120 (64.5%) | 0.07 |
| **AMR** |  |  |  |  |  |  |
| N | 87 | 347 |  | - | - | - |
| Age, mean (sd) | 59.3 (10.5) | 57.8 (11.5) | 0.29 | - | - | - |
| Female, N (%) | 29 (33.3%) | 175 (50.4%) | 6.00E-03 | - | - | - |
| **EAS** |  |  |  |  |  |  |
| N | 88 | 351 |  | - | - | - |
| Age, mean (sd) | 62.0 (10.4) | 51.2 (14.1) | 1.10E-09 | - | - | - |
| Female, N (%) | 33 (37.5%) | 215 (61.2%) | 9.60E-05 | - | - | - |
| **EUR** |  |  |  |  |  |  |
| N | 921 | 3656 |  | 262 | 714 |  |
| Age, mean (sd) | 62.8 (9.7) | 62.8 (9.7) | 0.96 | 61.3 (9.5) | 54.8 (11.6) | 8.10E-09 |
| Female, N (%) | 375 (40.7%) | 1498 (40.9%) | 0.88 | 111 (42.3%) | 472 (66.1%) | 3.40E-11 |
| **Combined** |  |  |  |  |  |  |
| N | 1195 | 4703 |  | 307 | 900 |  |
| Age, mean (sd) | 62.3 (9.8) | 61.1 (10.7) | 1.00E-03 | 61.3 (9.3) | 59.1 (13.1) | 0.04 |
| Female, N (%) | 499 (41.7%) | 2135 (45.4%) | 0.02 | 133 (43.3) | 592 (65.7%) | 6.40E-12 |

### GWAS identifies loci on chr 22q12 and chr19p13 as associated with risk of DME

There was no evidence of genomic inflation in this meta-analysis (λ_gc_ = 0.98 **Supplemental Figure 2**). We estimated the SNP heritability (h^2^) in the largest two super-populations, EUR and AFR, to be 0.41 and 0.48 respectively.

After genome-wide removal of non-DME specific loci via reverse regression, we found two novel loci at chr22q12.3 and chr19p13.11 reaching the pre-defined genome-wide significance level of P<5×10^−8^ (**Figure 1A** and **Table 2;** super-population specific results - **Supplemental Figures 3A-D)**, (Locus specific plots at **Figure 1B-C** (chr19p13.11, chr22q12.3 respectively) and super-population specific results - **Supplemental Figures 4A-H**).

**Figure 1:**
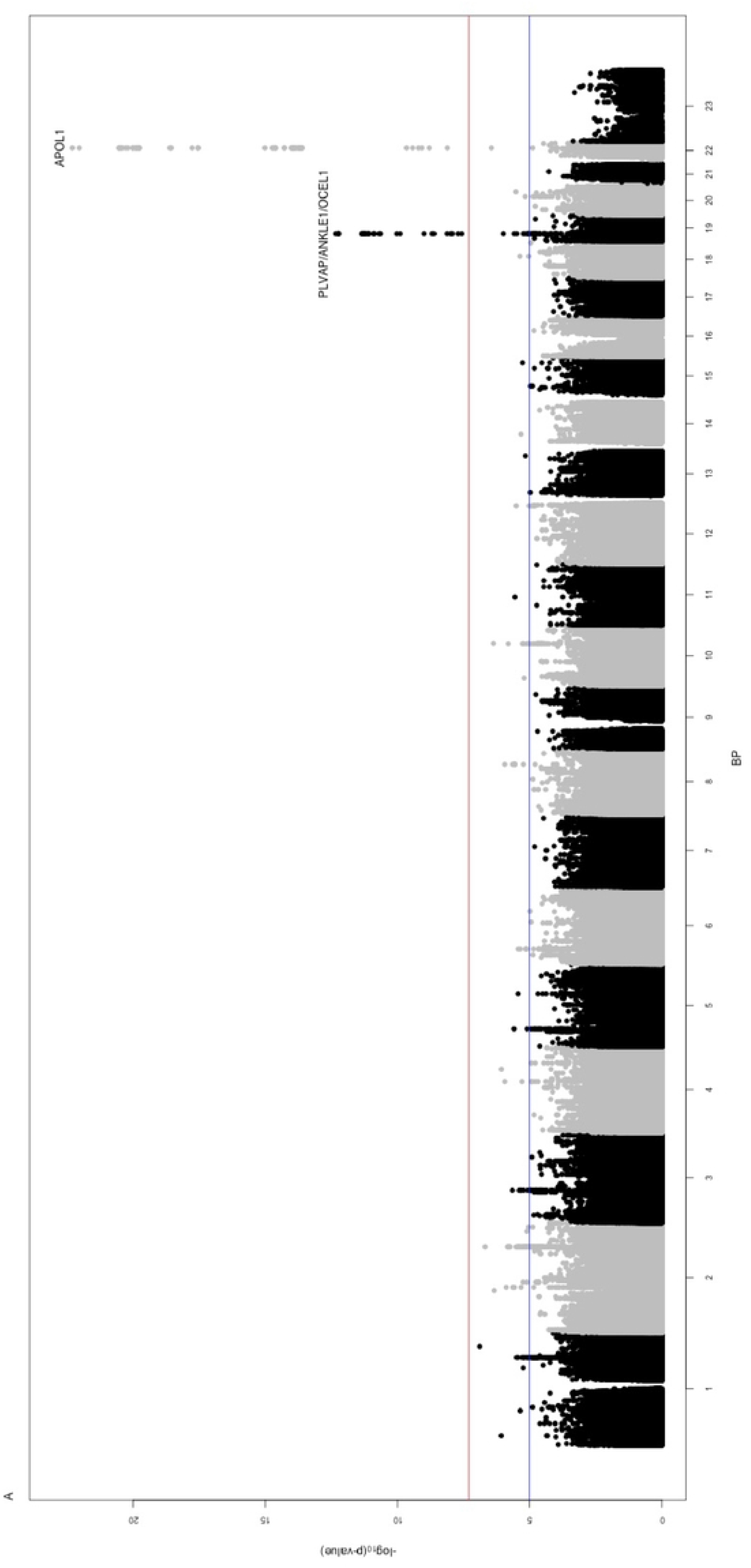
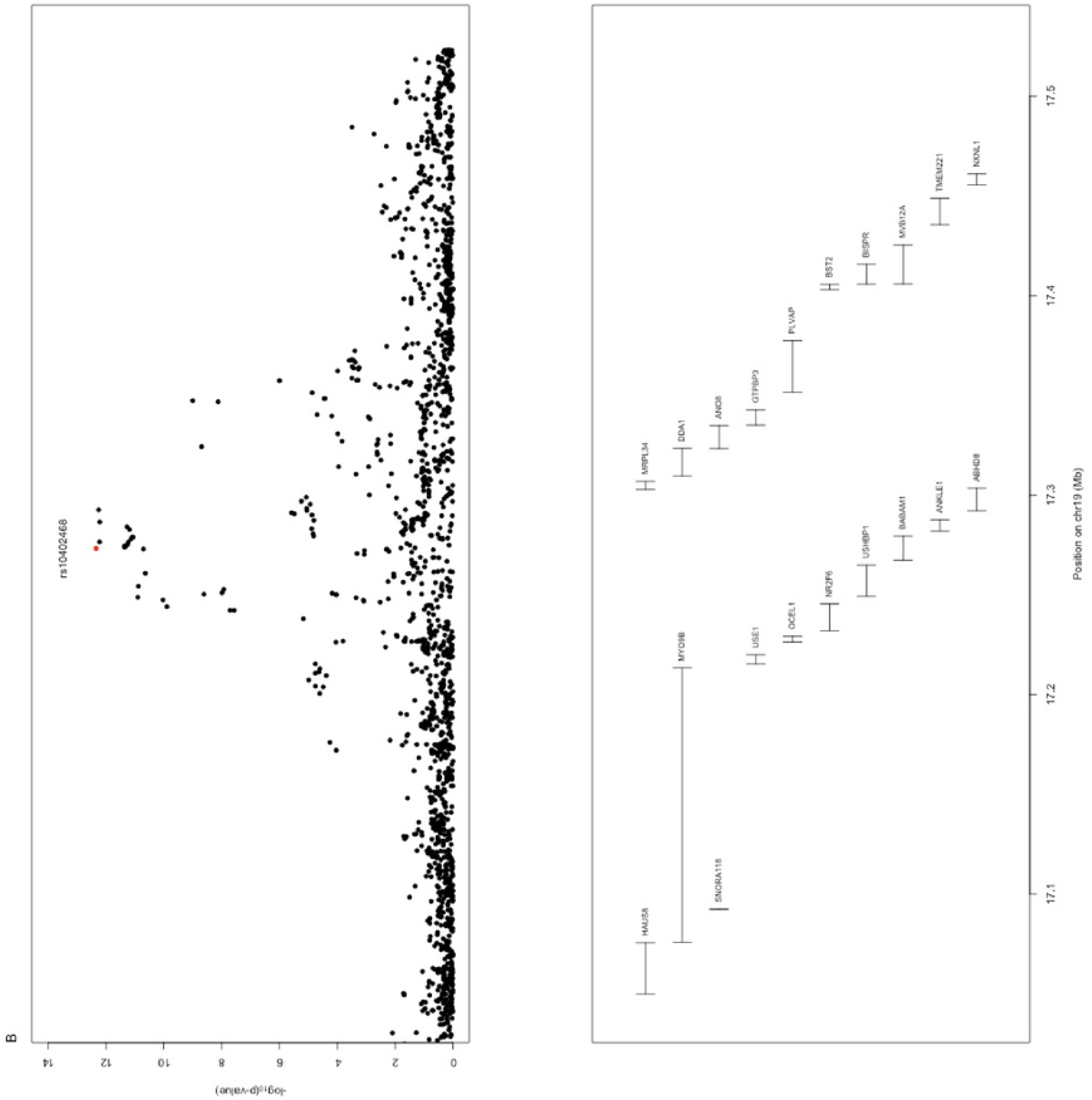
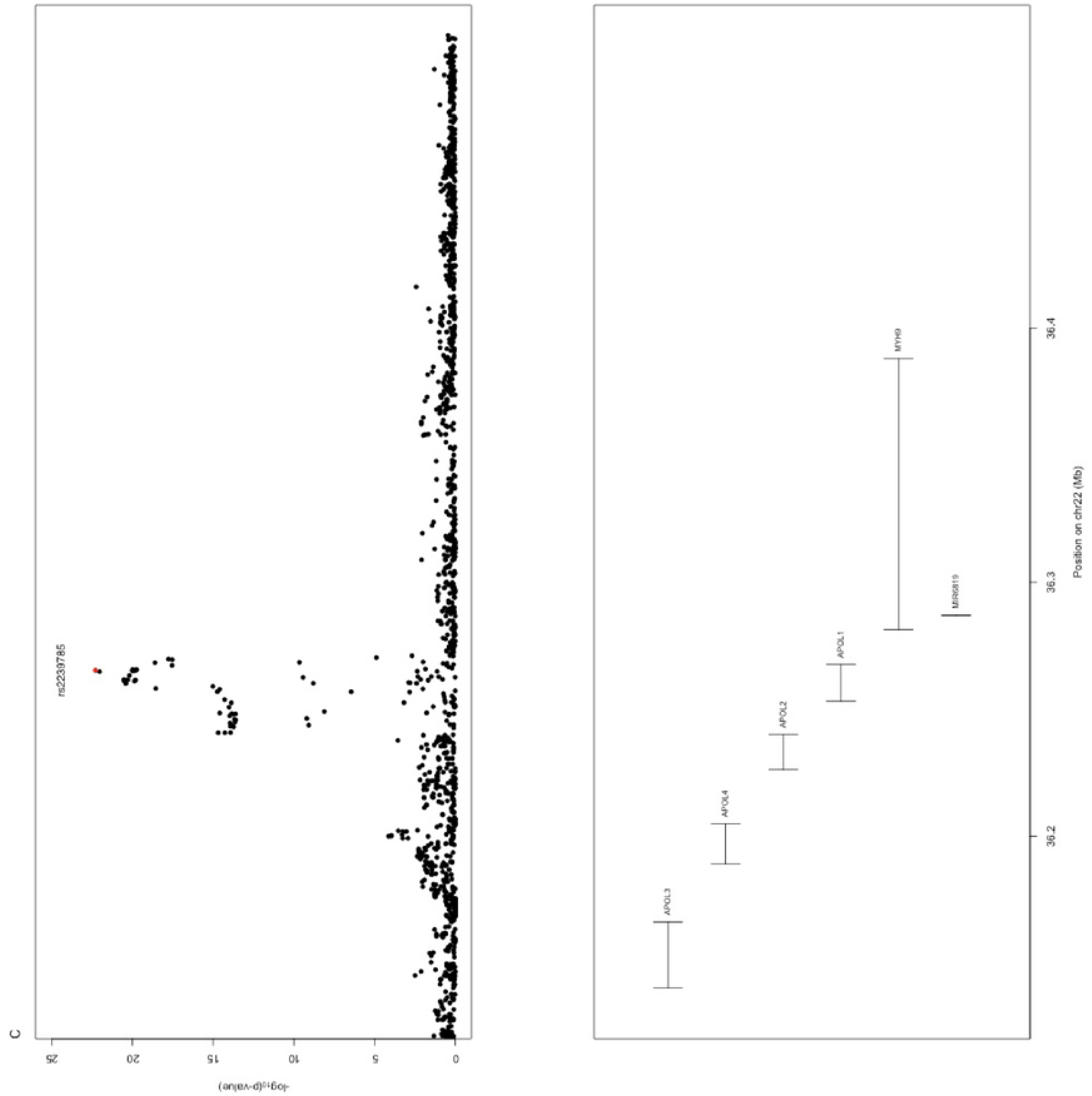
A) Manhattan plot of the discovery study population containing 1,195 DME cases and 4,703 non-DME controls. Gene names in peaks containing SNPs with a P<5×10^−8^ are labeled. The red line signifies the threshold for genome-wide significance (P<5×10^−8^) and the blue line indicates the threshold for suggestive significance (P<1×10^−5^) B) Regional plot (meta-analysis) for chr19. C) Regional plot (meta-analysis) for chr22. As the meta-analysis contains patient DNA samples from multiple ancestries, linkage disequilibrium information is not detailed here.

**Table 2.**
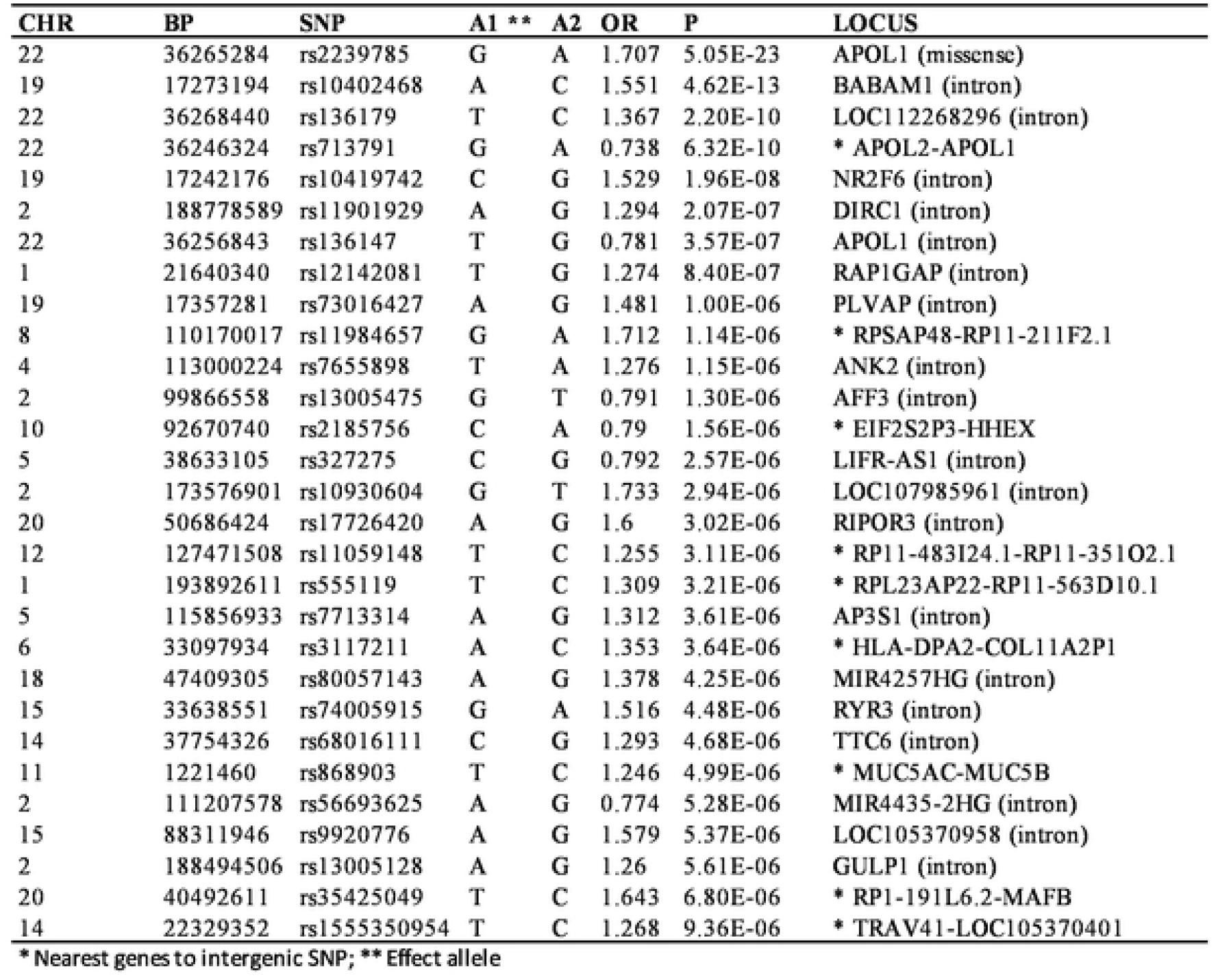
Independent list of SNPs that pass the suggestive threshold (meta-analysis).

### Chr22q12 – APOL1

The strongest signal of association in the discovery study population was at chr22q12.3 (**Supplemental Table 1**). The top SNP at this locus, rs2239785, results in a missense variant (K150E) in *APOL1*, encoding apolipoprotein L1 (G allele - OR_meta_= 1.71, P_meta_=5.05×10^−23^). The risk (G) allele encodes the E150 allele of the peptide. Previous investigation has shown the SNP is functional, with the E150 allele increasing APOL1-induced cytotoxicity in HEK293 cells compared to K150 expressed at similar levels(20).

The direction of effect was consistent across all four super-populations (OR_eur_=1.73, P_eur_=4.07×10^−20^; OR_afr_=1.84, P_afr_=0.002; OR_amr_=1.43, P_amr_=0.14; OR_eas_=1.43, P_eas_=0.12) (**Figure 2**). The frequency of this allele was highest in AFR DME patients (EAF_afr_ = 0.76), greater than twice that of EUR DME patients (EAF_eur_=0.31). We replicated the association at rs2239785 in an independent multi-ancestry DME case (PAGODA) and control study sample (OR_meta_=1.41, P_meta_=0.003; OR_eur_=1.31, P_eur_=0.03; OR_afr_=2.06, P_afr_=0.02), with a combined sample OR=1.65 and P=2.0×10^−24^ (**Table 3**).

**Figure 2:**
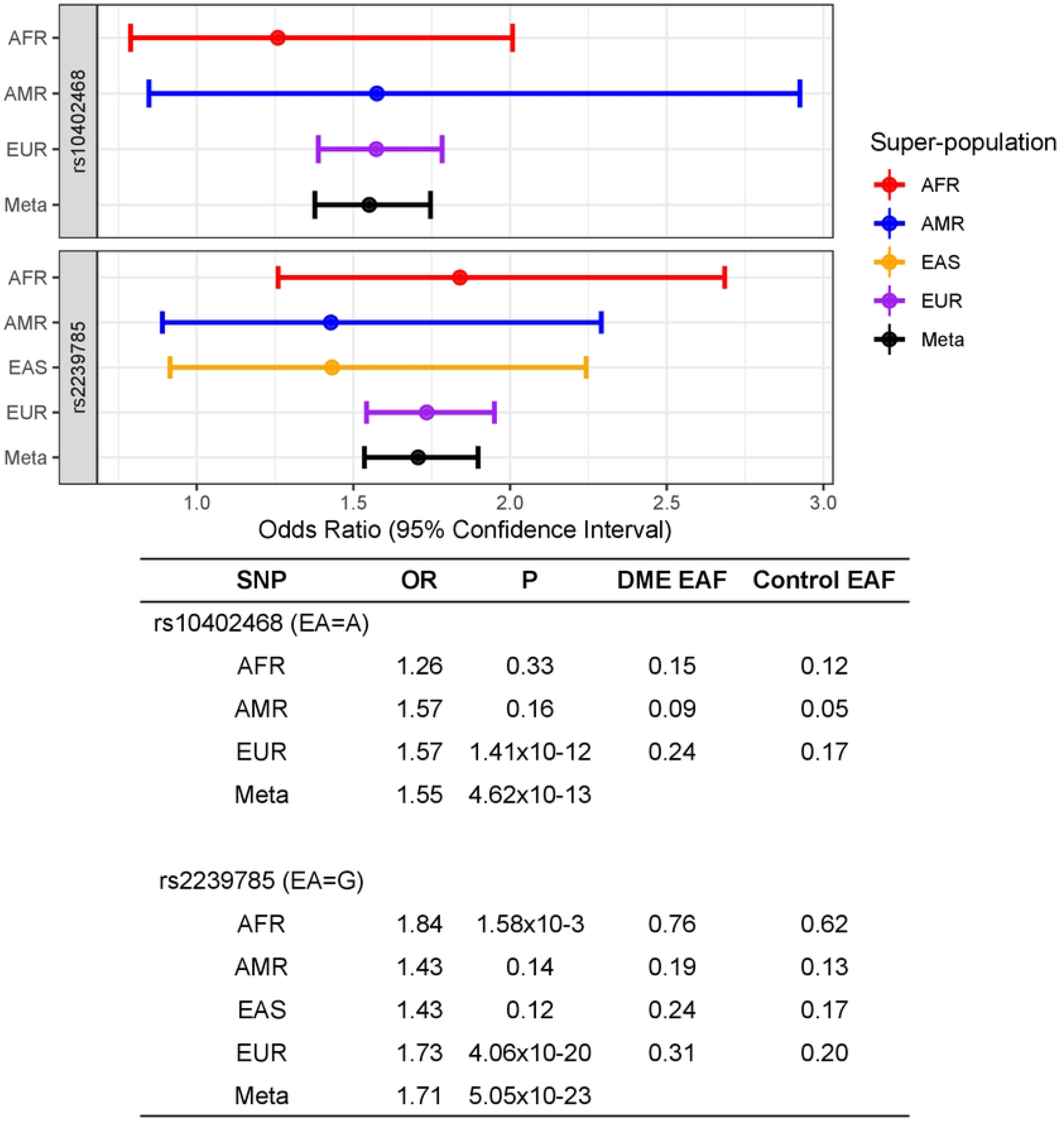
Forest plot and summary statistics for the top SNP at chr 22 and chr 19 loci.

**Table 3.** Replication of independent genom-wide significant hits.

| SNP | CHR | BP | A1 | A2 | Discovery<br>OR (P) | Replication<br>OR (P) | Meta-Analysis<br>OR (P) |
| --- | --- | --- | --- | --- | --- | --- | --- |
| rs10402468 | 19 | 17273194 | A | C | 1.55 (4.6E-13) | 1.31 (0.03) | 1.50 (1E-13) |
| rs2239785 | 22 | 36265284 | G | A | 1.71 (5E-23) | 1.41 (3.0E-3) | 1.65 (2E-24) |

To further investigate this association, we performed a PheWAS for rs2239785 in the FinnGen biobank study population (Freeze 7, https://r7.finngen.fi). FinnGen is a large biobank derived study where case assignment is based on physician coding, and cases were compared to the reminder of FinnGen study participants. In FinnGen, diabetic maculopathy was the only coded indication specific to diabetic complications of the macula, the area of the retina impacted in DME. For the index SNP at the *APOL1* locus found in this GWAS (rs2239785), we saw an association with diabetic maculopathy with the same direction of effect as the primary DME GWAS (β=0.14, P=5.5×10^−5^, MAF_cases_=0.18, MAF_controls_=0.16; N= 2,790 cases, 284,826 controls).

This was the second top association in all coded phenotypes for this variant, with the first being intrahepatic cholestasis of pregnancy (**Supplemental Table 2**). Finally we queried the summary statistics for a recent DME GWAS in Australian patients of EUR ancestry(21). While rs2239785 was not in the available data, we find a perfect proxy in rs136175 (r^2^=1.0, D’=1.0 with rs2239785 in EUR) has a similar direction of effect (OR=1.32, P=0.09; N=270 cases, 435 controls). Therefore, we are able to replicate the observed association between DME (or closely related phenotypes) and rs2239785, an amino acid changing SNP in *APOL1*, in three further studies.

Conditional analysis did not identify secondary signals at the locus in EUR; however SNP rs6518994 retained significance in the AFR sample after conditioning on rs2239785 in the primary GWAS dataset (original OR=1.92, P=2×10^−4^; conditioned OR=1.74, P=0.002; r^2^ with rs2239785=0.04). Further conditional analysis incorporating rs6518994 did not reveal any novel signals with P<0.01.

Due to the use of non-DME controls in this study for whom diabetes was not ascertained, it was important to establish whether the association we observed was driven diabetes status, rather than DME itself. We employed multiple approaches to exclude this. First, in the FinnGen PheWAS, we did not observe association with rs2239785 and T1D or T2D status (both P>0.53). Furthermore, we queried recent highly powered GWAS for type-1 (T1D)(22) and a multi-ancestry type-2 (T2D) diabetes finding no association for rs2239785 in either (both P>0.4). Further investigation in the EUR and AFR patient subsets of the T2D multi-ancestry study did not alter this (both P>0.5). Thus, we find this association is not driven by diabetes predisposition itself.

Multiple variants in *APOL1*, including rs2239785, have been associated with diabetic and non-diabetic nephropathy in African American individuals (23-28). These findings identified an AFR specific haplotype tagged by two SNPs (rs73885319, encoding S342G, and rs60910145, encoding I284M (G1)) and a 6-bp deletion (rs75781853 (G2)) associated with disease risk(23). G1 and G2, found in AFR only, are not strongly correlated (r^2^<0.18) but reside on the same haplotype block (D’=1.0) with rs2239785. Due to this haplotypic structure, it is difficult to disentangle the effect of rs2239785, G1 and G2 in AFR, and raises the possibility that the DME association we observed could be confounded to some degree by nephropathy. The G1 and G2 *APOL1* variants are not found in individuals with European ancestry. As such, the European population enables investigation of rs2239785 (K150E) on its own, and whether this SNP affects kidney disease.

First, we see repeated evidence of association between rs2239785 and risk of DME in individuals of European descent: the RISE, RIDE, YOSEMITE and RHINE trials in the discovery dataset, the PAGODA trial in the replication dataset and the FinnGen BioBank dataset. Second, in FinnGen, despite the association between rs2239785 and diabetic maculopathy, we found no evidence of association between rs2239785 and renal dysfunction in T1D or T2D patients (both P>0.37). Furthermore, we were unable to find an association between this SNP and DKD in recent GWAS conducted in European ancestry patients with either T1D or T2D (both P>0.71)(29; 30). Additionally, we saw no association in a large meta-analysis for CKD and kidney function (BUN, eGFR) in European patients (all P>0.22)(31). While renal failure or anticipated need for dialysis were exclusion criteria in the DME clinical trials from which these cases were obtained, a DKD diagnosis was not. There were kidney measurements for 912 of the DME patients in the YOSEMITE and RHINE trials. As a final test, we found that removing 290 patients from the 912 of who had albumin to creatinine ratio, blood urea nitrogen (BUN) or protein levels outside of the ideal range had no effect on the association with DME in the remaining 622 patients with normal kidney function (OR_meta_=1.79, P_meta_=7.91×10^−17^; OR_eur_=1.89, P_eur_=1.25×10^−16^; OR_afr_=1.76, P_afr_=0.03).

These data are consistent with a model whereby the G1 and G2 *APOL1* variants drive DKD susceptibility in individuals of African descent, but that rs2239785 (or another allele with which it is in linkage disequilibrium) represents an independent, and distinct, association that contributes directly to DME predisposition across ancestries.

### Chr19p13 – PLVAP and ANKLE1

The second genome-wide significant peak in this meta-analysis was located at chr19p13.11 (**Supplemental Table 3**) with the top SNP, rs10402468, located within intron 3 of *BABAM1* (A allele - OR_meta_= 1.55, P_meta_=4.62×10^−13^). The effect size and direction was similar across the super-populations tested (OR_eur_=1.57, P_eur_=1.42×10^−12^; OR_afr_=1.26, P_afr_=0.33; OR_amr_=1.58, P_amr_=0.15; monomorphic in EAS) (**Figure 2**). The risk allele was most common in EUR (EAF_eur_=0.25). We were able to replicate rs10402468 in an independent set of DME cases from the PAGODA trial and non-DME controls (OR_meta_=1.31, P_meta_=0.03; OR_eur_=1.34, P_eur_=0.03; OR_afr_=1.10, P_afr_=0.79) (**Table 1**), with a combined discovery and replication OR=1.50 and P=1.0×10^−13^ (**Table 3**). Conditional analysis did not identify a secondary signal at this locus. We derived the 95% credible set of variants for fine-mapping of the signal, identifying 4 potentially causal variants at the locus with 95% probability (**Supplemental Table 4**). Finally, we performed PheWAS in the aforementioned FinnGen study. While there was no association for diabetic maculopathy (P=0.11), there were several associations with ocular phenotypes when examining the locus at chr19p13.11 (rs10402468). The top phenotype in the PheWAS is macular cyst (β=0.35, P=4.6×10^−5^, MAF_cases_=0.31, MAF_controls_=0.23; N= 297 cases, 284,826 controls), and there are also strong associations with wet age-related macular degeneration (AMD) and “degeneration of macula and posterior pole” (P=1.3×10-4 and P=2.1×10^−4^ respectively) **(Supplemental Table 5)**. The AMD association did not replicate in a recent AMD GWAS (P=0.25; wet AMD vs controls) (32).

To investigate the possibility that this locus could be associated with diabetes status rather than DME we performed similar analyses as described for the locus at chr22. First, in the FinnGen PheWAS, we did not observe an association with T1D or T2D (both P>0.42). Second, there was no association for rs10402468 in large T1D(22) or T2D(33) GWAS studies (both P>0.3). Further investigation in T2D in the EUR patient subset of the multi-ancestry study did not alter this (P>0.4). Thus, we find this association is specific to DME, and not due to diabetes status.

This SNP, rs10402468, is in strong LD (r^2^=1.0) with a missense SNP in *ANKLE1* in all super-populations (rs10425939, A505V). We found that the peak co-localized with cis-eQTL peaks from bulk RNA-seq data(16-18) to *PLVAP* and *ANKLE1* in vascular/endothelium tissues (*ANKLE1* Prob_coloc_ artery aorta = 0.96; *PLVAP* artery coronary = 0.88, artery tibial = 0.60) (**Supplemental Figures 5A-C**). Thus, we identify both *PLVAP*, encoding plasmalemma vesicle associated protein, and *ANKLE1*, encoding ankyrin repeat and LEM domain-containing protein 1 as candidate effectors at the locus.

### Population attributable fraction at the APOL1 locus in AFR

The increased effect allele frequency of rs2239785 in AFR indicates a greater overall burden in this patient population. As such, we calculated the fraction of DME risk in patients with diabetes that is attributable to the *APOL1* locus **(Table 4)**. DME prevalence in United States patients with diabetes for non-Hispanic white and non-Hispanic black individuals is estimated at 2.6% and 8.4% respectively (3). We estimate that rs2239785 accounts for 5.2% of the risk of developing DME in non-Hispanic black patients with diabetes compared to 1.9% in non-Hispanic white patients. As previously mentioned, the G1 and G2 variants are only seen in AFR individuals and are on the same haplotype as rs2239785. We hypothesized that the population attributable fraction for rs2239785 in AFR who do not possess either the G1 or G2 haplotypes would be similar to that of EUR. Looking at this subsample in AFR, we estimate that rs2239785 accounts for 1.60% of the risk of developing DME in non-Hispanic black patients with diabetes. Thus, it appears the haplotype containing the rs2239785 risk allele (E150) itself, without the presence of G1 and G2 in AFR (most likely a result of recombination with the European rs2239785-only haplotype in the admixed African American population used in this study), accounts for a similar proportion of overall population risk in individuals of EUR and AFR ancestry. This provides further evidence that rs2239785 is a separate, DME specific locus which increases risk of DME regardless of presence of G1 and G2 in patients of African ancestry.

**Table 4.** Population attributable fraction for chr22 and chr19 loci by ancestry.

| Locus / Haplotype | Cohort | Population Attributable Risk Percent |
| --- | --- | --- |
| rs2239785 | EUR | 1.85 |
| rs2239785 | AFR | 5.16 |
| rs10402468 | EUR | 1.03 |
| rs10402468 | AFR | 0.74 |
| GG (EIK) Haplotype | AFR | 4.35 |

**Table 5.** DME Polygenic Risk Score In replication cohort.

| Population | PRS Cutoff | OR | P |
| --- | --- | --- | --- |
| AFR+EUR | 1.00E-05 | 1.36 | 2.80E-16 |
| AFR | 1.00E-05 | 1.17 | 0.253 |
| EUR | 1.00E-05 | 2.15 | 3.70E-29 |
| AFR+EUR | 5.00E-08 | 1.45 | 2.30E-03 |
| AFR | 5.00E-08 | 2.39 | 2.70E-02 |
| EUR | 5.00E-08 | 1.46 | 4.60E-03 |

### Association with baseline DME clinical and imaging measures

We sought to determine if these two loci impact DME clinical and imaging measures at baseline. We did not see an association for the *APOL1* and *ANKLE1/PLVAP* SNPs with baseline Diabetic Retinopathy Severity Score (DRSS) (N=1,168; P>0.1). Nor was there any association of the *APOL1* SNP with three retinal measures derived from Optical Coherence Tomography (OCT) data: (1) center subfield thickness (CST) (N=1,339), (2) intraretinal fluid in central subfield (N=1,020), and (3) subretinal fluid (N=387). However, we did observe an association between CST and the *ANKLE1/PLVAP* SNP (β=19.3; P=0.006). Thus, it appears the association of the risk allele with greater CST is consistent with previous reports of CST being higher in severe DME patients as well as when comparing DME patients to controls(34-36).

## Discussion

Despite multiple GWAS for DR and subtypes performed to date, few replicable associations have been reported. In this analysis we focused on the DME subtype to increase power by reducing disease heterogeneity.

In this multi-ethnic common variant GWAS, we report two novel loci for risk of DME. Both loci achieve genome-wide significance in the discovery dataset and replication in both an additional multi-ancestry DME clinical trial patient sample, and the FinnGen biobank study. We observed consistent effect sizes and directions across ancestries in both the discovery and replication datasets at both loci.

We also observed super-population differences in disease heritability, where the SNP-heritability was 0.41 in EUR and 0.48 in AFR. This was partially due to variation at the *APOL1* locus in AFR, where the effect allele frequency for the top SNP was over two times that of EUR. This increased allele frequency at the *APOL1* locus resulted in an estimated 5% population attributable fraction of DME in non-Hispanic black patients with diabetes, over two times that of non-Hispanic white patients, despite the similar effect size for the SNP in each super-population. This indicates that non-Hispanic black patients with DME could preferentially benefit from therapies designed around APOL1.

To date there are no published reports of APOL1 staining in the eye, but scRNA seq data indicates expression in immune and vascular cells of the retina and choroid endothelia(37). The development of macular edema requires a greater rate of fluid entry from leaky vessels into the retina than that of fluid reabsorption(38). Given that *APOL1* renal risk variants expressed under an endothelial promoter in mice caused vascular leak in the skin(39), one possible mechanism is that APOL1 could similarly exacerbate vascular leak in the retinal or choroidal endothelia, directly resulting in edema. Alternatively, the leaky vasculature could allow infiltration of cytokines into the eye, which would upregulate APOL1 beyond a critical threshold in the cells where it is expressed, causing cytotoxicity(40). Another possibility is that the cation channel activity of APOL1 (e.g. K^+^ efflux) could interfere with K^+^ co-transporter mediated water reabsorption by Müller or retinal pigment epithelial cells, contributing to edema(38; 41). Determining the localization of APOL1 protein in DME eyes would help narrow down the possibilities.

The second locus identified includes the genes *PLVAP* and *ANKLE1*. PLVAP, plasmalemma vesicle-associated protein, is also called PV-1 and recognized by the PAL-E antibody. PLVAP is an endothelial protein that regulates basal permeability. In the eye, PAL-E staining is specific to the fenestrated endothelium of the choriocapillaris and ciliary processes(42). *Plvap* deficient mice develop edema in some vascular beds(43) and loss of Plvap has been shown to disrupt the choriocapillaris structure and lead to retinal degeneration(44). As such, these murine studies are consistent with our results suggesting lower *PLVAP* mRNA levels are a risk factor for DME. However, *PLVAP* expression is also seen at the blood retinal barrier (BRB), but only after insult by factors that induce leakage, such as diabetic retinopathy(45). In a properly functioning BRB, *PLVAP* expression is low or non-existent(46). This is contrary to our results and further studies will be needed to associate the risk allele with differential PLVAP expression and function.

*ANKLE1* (which encodes the protein LEM3) was also identified at this locus by our co-localization analysis. LEM domain proteins, such as LEM3 bind BAF, a chromatin protein. ANKLE1 has been shown in *C. elegans* to be important in the formation and stabilization of chromatin bridges in the end stages of cellular division(47). Expression appears somewhat ubiquitous with enrichment in brain and eye tissue(48), however mechanistic information on ANKLE1 and retina or eye is absent from the literature. While further perturbation studies will be required to assess causality, based on what is currently known, *PLVAP* is the most likely causal gene at this locus.

Notably, we observed no prior DME, DR or PDR loci at the GWAS significant level in this DME analysis (**Supplemental Table 6**)(8; 10; 21). This could be for multiple reasons. First, this study contains at least three times more cases than previous DME studies. Additionally, other studies have focused on DR patients as a whole. As DR is a heterogeneous condition, homogenizing based on phenotypes within DR can increase power to find an association.

A weakness of our study is the use of controls from non-ophthalmologic clinical trials. This has the potential to lead to associations that are confounded by diabetes status or other diabetic complications such as kidney disease. This is especially important at the *APOL1* locus, for which multiple alleles in *APOL1* have strong associations to risk of kidney disease in individuals of African descent. We have addressed this in several ways, including leveraging genetic architecture differences at the *APOL1* locus in individuals of European and African descent. We queried highly powered GWAS for T1D and T2D, finding no hint of an association for either locus in patients of European ancestry. For the *APOL1* locus we queried DKD, CKD and kidney function GWAS, again finding no association in European ancestry patients. Through PheWAS in the FinnGen population, we found strong associations for ocular complications due to diabetes, but not T1D, T2D or kidney disease. Finally, we conducted a sensitivity analysis, where we removed patients with signs of kidney malfunction at baseline finding no effect on the association at the *APOL1* locus. Thus, these findings appear to be specific for DME risk in multiple populations and not related to the concomitant diagnosis of diabetes or other diabetic complications. Historically, the standard design when assessing determinants of complications from diabetes has been to compare patients with complications from diabetes to patients with diabetes and no complications. Our findings question the need for such a constraint on study design, especially when considering there are multiple highly powered GWAS one can query to detect possible confounding due to diabetes risk itself or other co-morbid complications.

In summary, here we present results from the largest GWAS on DME to date which highlights *APOL1*, a gene canonically associated with kidney disease, and broadens its impact to a second diabetic complication. We find this gene to be associated with risk of DME in multiple populations, and note a substantial increased genetic risk of DME in non-Hispanic black patients with diabetes due to the increased allele frequency of rs2239785 in patients of African ancestry. Additionally, we uncovered an additional locus containing the genes *ANKLE1* and *PLVAP*. Historically, replicable loci for diabetic retinopathy and sub-phenotypes have been difficult to uncover. The present study indicates that power to detect genetic effects influencing diabetic ocular phenotypes can be improved by categorizing DR cases by sub-phenotype prior to genetic analysis.

## Data Availability

We are able to provide summary statistics for the genetics analysis upon request to the Corresponding Author (BLY). Interested researchers should contact the Corresponding Author for information on how to access the complete summary statistics for this manuscript.

## Acknowledgements

We would like to acknowledge Hsu-Hsin Chen and Luz Orozco for their efforts to identify APOL1 expression in retina.

We would like to acknowledge the efforts of Marie Waldvogel, Aurelie Felez and Nicole Bienz in the Pharma Biosample Services (PBS) Department of F. Hoffmann-La Roche for their efforts on handling and genotyping the patient samples used in this study.

We also would like to acknowledge the participants and investigators of FinnGen study. The FinnGen project is funded by two grants from Business Finland (HUS 4685/31/2016 and UH 4386/31/2016) and the following industry partners: AbbVie Inc., AstraZeneca UK Ltd, Biogen MA Inc., Bristol Myers Squibb (and Celgene Corporation & Celgene International II Sàrl), Genentech Inc., Merck Sharp & Dohme Corp, Pfizer Inc., GlaxoSmithKline Intellectual Property Development Ltd., Sanofi US Services Inc., Maze Therapeutics Inc., Janssen Biotech Inc, Novartis AG, and Boehringer Ingelheim. Following biobanks are acknowledged for delivering biobank samples to FinnGen: Auria Biobank (www.auria.fi/biopankki), THL Biobank (www.thl.fi/biobank), Helsinki Biobank (www.helsinginbiopankki.fi), Biobank Borealis of Northern Finland (https://www.ppshp.fi/Tutkimus-ja-opetus/Biopankki/Pages/Biobank-Borealis-briefly-in-English.aspx), Finnish Clinical Biobank Tampere (www.tays.fi/en-US/Research_and_development/Finnish_Clinical_Biobank_Tampere), Biobank of Eastern Finland (www.ita-suomenbiopankki.fi/en), Central Finland Biobank (www.ksshp.fi/fi-FI/Potilaalle/Biopankki), Finnish Red Cross Blood Service Biobank (www.veripalvelu.fi/verenluovutus/biopankkitoiminta) and Terveystalo Biobank (www.terveystalo.com/fi/Yritystietoa/Terveystalo-Biopankki/Biopankki/). All Finnish Biobanks are members of BBMRI.fi infrastructure (www.bbmri.fi). Finnish Biobank Cooperative -FINBB (https://finbb.fi/) is the coordinator of BBMRI-ERIC operations in Finland. The Finnish biobank data can be accessed through the Fingenious® services (https://site.fingenious.fi/en/) managed by FINBB.

## Author contribution statement

ADS and BLY conceived and designed the study. ADS did the statistical analysis. MCC, WF, NA, AM, RP, MIMcC, and SJS contributed to the experimental design and analysis. SS, VAI, JR, EW and NMC contributed to whole-genome sequence generation, rGWAS chip data generation, single nucleotide polymorphism calling, and analysis pipeline generation. LM, ANN, and MPvdB were site principal investigators responsible for participant recruitment and data collection. ADS and BLY wrote the first draft of the manuscript, which was reviewed by all authors.

## Code Availability

The code that supports the findings of this study are available on request from the corresponding author (BLY).

